# Perceived mental health, wellbeing and associated factors among Nepali migrant and non-migrant workers: A qualitative study

**DOI:** 10.1101/2020.11.23.20236794

**Authors:** Hridaya Raj Devkota, Bishnu Bhandari, Pratik Adhikary

**Affiliations:** Institute for Social and Environmental Research Nepal (ISER-N); Community Support Association of Nepal (COSAN), Kathmandu, Nepal; Health Research and Development Forum (HRDF), Nepal; School of Public Health, UC Berkeley, USA

**Keywords:** Migrant workers, Mental health, Wellbeing, Affecting factors, Labor workers, Nepal

## Abstract

**Background:** Poor mental health and illness among the working population have serious socio-economic and public health consequences for both the individual and society/country. With a dramatic increase in work migration over the past decades, there is recent concern about the health and wellbeing of migrant workers and their accessibility to healthcare services in destination countries. This study aimed to explore the mental health and wellbeing experiences of Nepali returnee-migrants and non-migrant workers, and identify their perception on the risk factors for poor health and health service accessibility for them.

**Methods:** This qualitative study was conducted among Nepali migrant and non-migrant workers in February 2020. Four focus group discussions (n=25) and 15 in-depth interviews were conducted with male non-migrant and returnee migrant workers from Gulf countries and Malaysia. The discussions and interviews were audio-recorded, transcribed, translated into English and analysed thematically.

**Result:** Migrant workers reported a higher risk of developing adverse mental health conditions than non-migrant workers. In addition, fever, upper respiratory infection, abdominal pain, ulcer, and occupational injuries were common health problems among both migrant and non-migrant workers. Other major illnesses reported by the migrant workers were heat burns and rashes, snake-bites, dengue, malaria, gallstone, kidney failure, and sexually transmitted diseases, while non-migrants reported hypertension, diabetes, and heart diseases. Adverse living and working conditions including exploitation and abuse by employers, lack of privacy and congested accommodation, language barriers, long hours’ hard physical work without breaks, and unhealthy lifestyles were the contributing factors to migrant workers’ poor mental and physical health. Both migrant and non-migrants reported poor compliance of job conditions and labor protection by their employers such as application of safety measures at work, provision of insurance and healthcare facilities that affected for their wellbeing negatively. Family problems compounded by constant financial burdens and unmet expectations were the most important factors linked with migrant workers’ poor mental health condition.

**Conclusion:** Both migrant and non-migrant workers experienced poor mental and physical health condition largely affected by their adverse living and working conditions, unmet familial and financial needs and adherence to unhealthy life styles. It is needed to ensure the compliance of work agreement by employers and promotion of labor rights in relation to worker’s health and safety. In addition, policy interventions on raising awareness on occupational health risk and effective safety training to all migrant and non-migrant workers are recommended.

## Background/Introduction

International labor migration is a growing phenomenon globally. It is estimated that 164 million people are migrant workers across the world with a rise of 9% from 2013 to 2017 [1]. The largest flow of migrant workers from South Asia is to Gulf Cooperation Council (GCC) countries and Malaysia. Nepal is one of the largest source countries in South Asia for labor supply to those countries [2].

In the last two decades, foreign employment has become attractive for Nepali working age population. Every year over 300,000 Nepali migrate abroad for employment, mostly to the Gulf countries and Malaysia. Over 85% of the total workforces migrated comprised largely semi-skilled having low educational background, predominately working in the construction, industry and service sectors. [2,3]. Labor migration has played an important role in the Nepalese economy. Remittance by migrant workers has contributed to over a quarter of the country’s GDP in recent years, which stood at 8.79 billion USD in 2018/19 [3].

Despite those benefits, there is serious concern about the mental health, wellbeing, and safety of the Nepali migrant workers due to the high number of injuries and deaths during their employment abroad. The data shows that the reported deaths among Nepali labor migrants abroad was 4320 for the period of FY 2008 – 09 to FY 2014 – 15. The report also shows that 21.8% of all deaths were due to cardiac arrest, and 10.4% suicide. Over 97% of those deaths occurred in GCC countries and Malaysia [4].

Migrant workers are usually employed in high risk and hazardous sectors such as construction, mining/ industries, factories, agriculture, and the service sector. These kinds of jobs typically involve long hours and hard physical labor, which can result in increased occupational accidents and illnesses [5,6]. They often work in poor conditions with low wages. Moreover, migrant workers experience abuse, exploitation, and psychosomatic effects of stress at work. Sexual exploitation has been commonly reported in the case of female labor migrants working in informal sectors, such as domestic work [7]. Previous studies showed that migrants who experienced workplace abuse, exploitation and perceived insecurity increased risk of mental illness [7,8]. Furthermore, cultural incompatibility, isolation, lack of social support and living away from the family were commonly reported risk factors for psychological distress among migrant workers [9,10].

In addition to occupational health vulnerabilities of migrant workers, they are prone to HIV, Tuberculosis, mental health illness due to their engagement with multiple sex partners, poor living condition and life style, and lower socio-economic status [11,12]. Previous studies also reported that migrant workers do not pay much attention to their health and seeking care unless they get serious problem due to financial difficulties, and also due to the risk of job termination and deportation (13,14). Furthermore, the lack of a sound health system and Nepal’s limited capacity to provide quality mandatory health examination prior to departure and having no provision of returnee health check-up services in the country upon arrival, Nepali migrant workers are more at risk of illness [4,7].

The literature also shows higher rate of occupational accidents, injuries and frequency of sicknesses among in-country Nepali workers due to unhealthy and hazardous workplaces with poor safety practices. Moreover, repetitive stress and other chronic diseases are also found common among them [15]. The nature and magnitude of the problems varied by the type of job, however, common illnesses that they suffered were diarrheal diseases, ENT problems, chest infection, joint and back pain. Besides, the construction workers suffered from gall-stone, heart diseases and urinary problems [15,16]. There is a limited information on mental health and wellbeing of Nepali migrant and non-migrant workers and their healthcare access in the country and abroad. This study aimed to explore the experience of and factors affected to mental health and wellbeing among Nepali migrant and non-migrant male workers. The study also assessed the common health problems encountered by those groups, and accessibility of healthcare services for them in Nepal and abroad.

## Methods

### Study design

A cross-sectional study using qualitative approaches to data collection was conducted in February 2020. Four focus group discussions (FGDs) and 15 in-depth interviews with non-migrants and returnee migrant workers having a range of socio-cultural and occupational backgrounds were conducted to understand the experience of study participants’ mental health and their views on the factors affecting their health and wellbeing.

### Study setting

The study was conducted in Madi Municipality of Chitwan, a southern district of Nepal with a population of 43,402 with 29% of males aged 19 – 59 years [17]. It is one of the fast growing urban municipalities with the influx of rural population from around the neighboring districts with heterogeneity of population characteristics in terms of caste, ethnicity, socio-cultural and economic status. In terms of caste breakdown by the 2011 census, the study area population comprised of 45% Janajati (indigenous), 35% Brahmin and Chhetri, 18% Dalit, and 2% others such as Madhesi and Muslims [17]. Municipality records show that a total of 2,627 males in foreign employment, mostly in the Gulf Countries and Malaysia [18].

### Study participants and recruitment

The study participants were purposively selected using specific criterion. Two categories of participants – non-migrants and returnee migrants, all males from range of social strata and occupational groups were recruited for interviews and FGDs. Of the total 40 participants (19 non-migrants and 21 returnee migrants), 15 were recruited for face-to face semi-structured interviews, while 25 participants were selected and recruited for FGDs. Two FGDs with non-migrants and two with returnee migrant workers consisting 6 - 7 participants in each group were conducted. Interviews and discussions were held in a natural setting. Semi-structured interviews were taken individually in their homes and FGDs were conducted in four different locations with non-migrant and returnee migrants’ groups. Participants from diverse social background were recruited to capture their views from multiple perspectives. The number of interviews and focus group discussions were determined by data saturation [19].

### Ethical consideration

The study protocol was approved by Nepal Health Research Council (NHRC) - Ref. No: 490. We also obtained approval from study site local authority for the study. In addition, written consent of each participant was taken prior to their interviews and discussions after describing the purpose of study, their voluntary participation, the confidentiality and anonymity of information that they provide.

### Data collection tools and procedure

The interview and FGD topic guides were developed and tested before use. The guides covered the questions related to participant’s perception and knowledge about mental health, experience on any mental health symptoms, other illnesses, and their opinion on the factors affecting their health. The topic guide also included questions related to healthcare access and quality of services that they received. Moreover, the tools incorporated questions about participant’s job nature, living and working environment and their individual behavior such as smoking, consuming alcohol and means of entertainment. The tools were developed, and the interviews were conducted in Nepali.

Two experienced interviewers in qualitative data collection with the help of two local trained research assistants, conducted the discussions and interviews. The role of the research assistant was to obtain consent of participants and to take notes during interviews and discussions. Developing a sustained contact, we fostered a relationship with study participants and encouraged their contribution. Considerable effort was put into maintaining neutrality and balancing the power relationship between the researcher and the participants at all stages of the research process. All interviews and discussions were audio-recorded with participant’s written approval.

### Data Analysis

After completion of field data collection, we followed a series of steps before the analysis proceeded to the interpretive phase. The first step involved transcribing verbatim all the audio-recordings in Nepali and translation into English which was done by the second author and two other language specialists. Then the first and last authors reviewed all transcripts and the interview notes, reading, rereading and reviewing for overall understanding. Following the framework method developed by Ritchie and Spenser, we then analysed data in five stages: familiarization; identifying a thematic framework; indexing; charting/mapping; and interpretation [19]. To ensure accuracy for inter-rater reliability, the last author who also assisted in conducting the interviews, crosschecked the transcriptions, translations and data coding. At this stage, where no new concepts emerged from the further review and coding of data, we developed sub-themes and grouped together the concepts identified in the text based on their similarities and relationships to develop themes and subthemes (Table 1). The themes and subthemes were then analyzed in relation to the research objectives and are described in the following section.

**Table 1:**
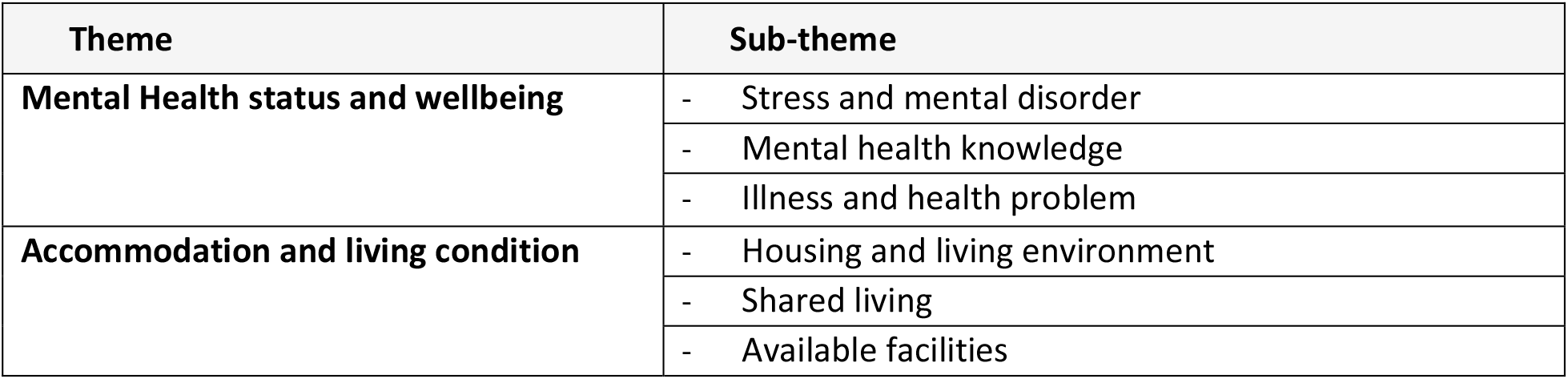
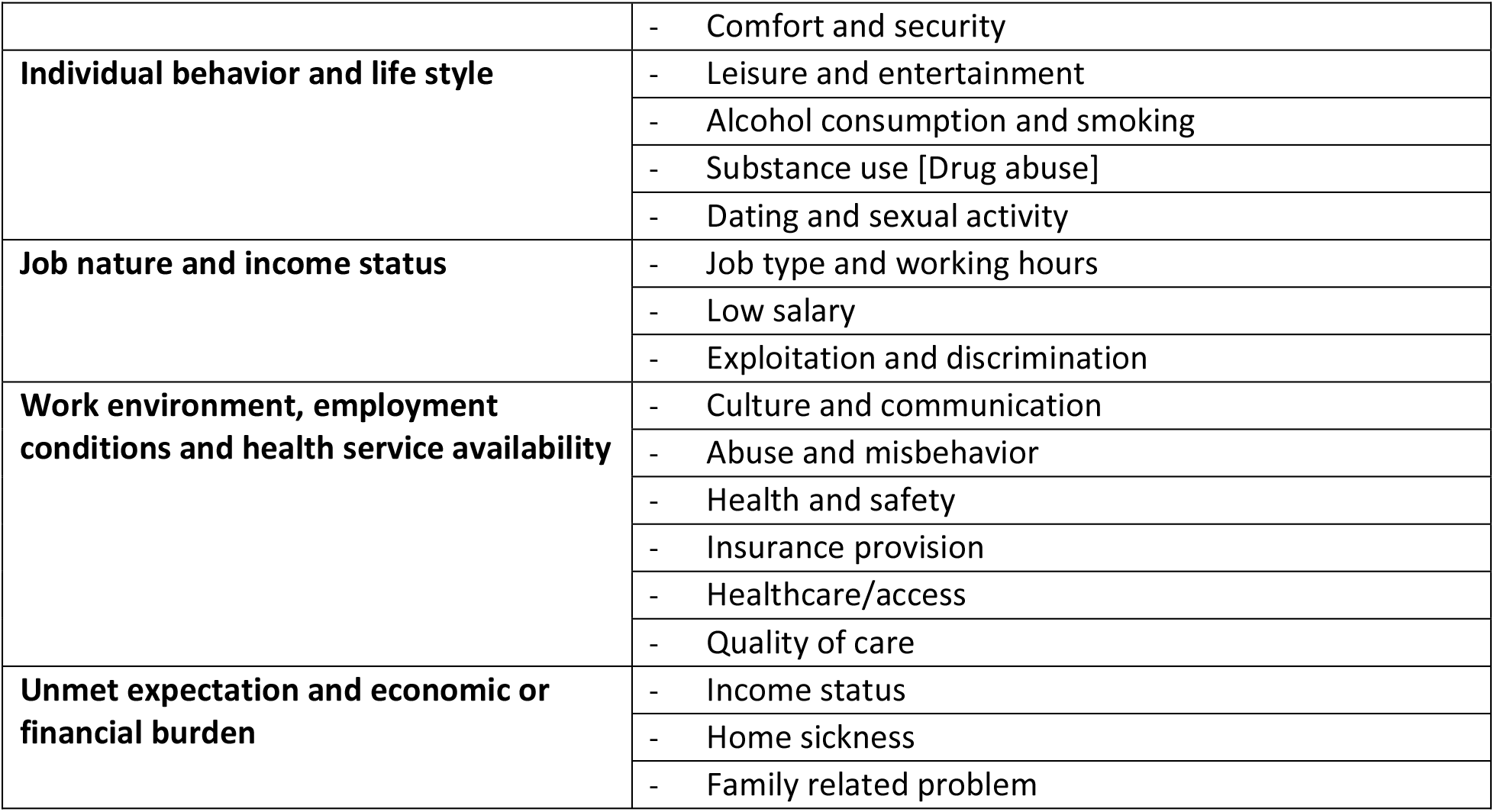
Themes and Sub-themes

| Theme | Sub-theme |
| --- | --- |
| **Mental Health status and wellbeing** | - Stress and mental disorder |
|  | - Mental health knowledge |
|  | - Illness and health problem |
| **Accommodation and living condition** | - Housing and living environment |
|  | - Shared living |
|  | - Available facilities |
|  | - Comfort and security |
| **Individual behavior and life style** | - Leisure and entertainment |
|  | - Alcohol consumption and smoking |
|  | - Substance use [Drug abuse] |
|  | - Dating and sexual activity |
| **Job nature and income status** | - Job type and working hours |
|  | - Low salary |
|  | - Exploitation and discrimination |
| **Work environment, employment conditions and health service availability** | - Culture and communication |
|  | - Abuse and misbehavior |
|  | - Health and safety |
|  | - Insurance provision |
|  | - Healthcare/access |
|  | - Quality of care |
| **Unmet expectation and economic or financial burden** | - Income status |
|  | - Home sickness |
|  | - Family related problem |

## Results

### Characteristics of study participants

Of the total 40 participants, 53% were returnee migrants almost from Gulf countries – Saudi Arabia, Kuwait, UAE and Qatar, and a few of them returned from Malaysia. The mean age of the participants was 34.6 (SD ± 9.7) ranging 19 – 55 years. 73% of study participants had secondary or higher level education, while less than 8% had primary level of education. The majority of participants were Jana Jaati (60%) followed by Brahmin and Chhetri (37.5%). A small proportion (7.5%) were Dalits. In terms of occupation of returnee migrants abroad, 48% worked as laborers, 19% as drivers, and 33% worked as semi-skilled labor, security guards and office boy/cleaners. Similarly, out of 19 non-migrant participants, the majority (53%) worked as laborers, over one-fifth worked in government or private offices as lower level staff or technicians, 16% worked in the service sector and 11% as primary school teachers.

### Mental health status and wellbeing

#### Stress and Mental Disorder

Both migrant and non-migrant participants in their focus group discussions and interviews stated that they often experienced various level of stress and mental sickness during their foreign employment. It was also reported that a few migrant workers returned back to Nepal terminating their employment because of sustained mental health issues. The participants also reported cases of suicide of others, which they believed was caused by severe stress and depression that they developed due to their adverse work environments and family problem.

> “There are many people experiencing tension and stress abroad. People are in double burden as they experience tension at work as well as from home. I myself cannot sleep well. The employer put on pressure even in a minor mistake at work.”
>
> IDI, Returnee-migrant
>
> “I have seen a number of Nepalese people in difficult and stressful situation abroad. One of my friends became alcoholic and often slept on road with the reason, as he shared that his wife got married with someone else back home and also took all the money that he earned. He didn’t want to return back to Nepal and often attempted suicide. We helped him and sent back to Nepal.”
>
> IDI, Returnee-migrant

Non-migrant participants expressed their mixed experience on the stress and mental disorder among themselves. Seasonal labor workers, and those who earned very little and not enough to meet their basic needs, reported living with stressful life. They further stated that even with long working hours, usually 10 – 12 hours daily, they earned very little due to the low pay rates in Nepal. However, those who had regular jobs, had their own farming land and produced agro-products did not report any symptoms of mental sickness. This group of non-migrant participants both in their interviews and FGDs reported their satisfaction with their employment, income and family life. In contrast those working in others stated:

> “Many people who don’t have job opportunity are stressed and suffering from depressive disorders. We have low salary so we often feel very stressed to manage the needs of family. Some employers or contractors don’t pay on time that creates more tension especially while the family member get sick and needed hospitalization.”
>
> FGD, Non-migrant
>
> “I often experienced unhappy mood, stress, tension, humiliated and sometimes not hoping to live. This happened while I faced economic challenges and not able to manage this. Due to the stress I had poor sleep.”
>
> IDI, Non-migrant

#### Mental Health Knowledge

When asking about the mental health knowledge, most of the interview participants could not clearly describe symptoms of mental illness. Also, many of them were not aware of the consequences of long-term stresses on the health. However, a few did have some knowledge about the symptoms of mental health problems, and also stated that it may lead on individual to suicide. Some others said that people have knowledge but they do not care and perceive it as a health problem.

> “I don’t have much idea. Usually, people don’t have knowledge about mental health. We often face tension or stress but we don’t take it as a health problem. Many don’t know about the negative consequences of tension and stress for health.
>
> FGD, Returnee-migrant

#### Other illness and Health Problems

In addition to mental distress, study participants reported that migrant workers often suffered from other health problems and illness. Commonly reported illness included seasonal flu, fever, cough, cold, tonsillitis, chest pain, respiratory diseases, abdominal pain and ulcer. Moreover, neck and back pain, injury and paralysis were reported mostly by labor workers involved in heavy lifting and construction works. Furthermore, participants returning from Gulf countries complained of heat burns and rashes, while those from the Malaysia reported snakebites, Dengue and Malaria. It was also reported that some of them or their friends suffered from gall-stones, kidney failure and cancer, and returned back to Nepal terminating their employment.

> “……of course, not only stress related…. we also suffered from other illness there. In the initial period, sore throat, tonsillitis and fever was more common among most workers, may be it was due to mismatch of weather and changed environment. ….and also other serious illness such as ulcer and kidney diseases. One friend returned Nepal due to his sickness.”
>
> IDI, Returnee-migrant

Common health problems reported by non-migrants were the common cold, fever, hypertension, diabetes and heart disease. Both returnee migrant and non-migrant workers commonly reported that they faced problems of occupational accidents and injuries. The intensity of this problem increased while working abroad.

> “There, I observed frequently the cases of accident and injuries. One of our storekeeper got an accident at work and he had leg fracture and ligament damage. The company provided full treatment and two months paid leave.”
>
> IDI, Returnee-migrant
>
> “Oh! I have witnessed an accident. One of my friends fell from the height while at work. He got back injury. We supported him to take hospital and he was recovered after treatment. However, the employer did not support him at all for his treatment.”
>
> IDI, Non-migrant

A few participants also reported that they suffered from sexually transmitted diseases as they involve in dating and unsafe sexual activities abroad. One of the interview participants shared that:

> “Um…, I suffered from a sexual problem abroad. I was involved in sexual activities after seven months while being abroad. After one and half months of sexual contact, I experienced pain and allergy in my penis. I got the treatment there and even after returning back I checked my blood and reported normal but I am still experiencing some problem.”
>
> IDI, Returnee-migrant

#### Factors associated with mental distress and wellbeing

Migrant workers reported a range of factors that affected their mental health and wellbeing. Key factors included: their accommodation and living condition, individual behavior and life style, job nature and income status, work environment and employment condition, unmet expectation and economic burden.

#### Accommodation and living condition

Migrant workers commonly reported that they had poor living conditions without basic facilities that often created tension and feeling of insecurity. Most of them reported their shared accommodation provided by the company while working abroad. A few of them reported that they had a reasonable accommodation with enough space and basic facilities, while most participants said the rooms were congested not having basic facilities such as beds, cabinets, refrigerators and kitchen amenities.

“We did not get apartment as we expected. We were 8 – 10 in our room, too crowded. We did not have any space to store our luggage’s, to put our clothes. Often we needed to find out some space outside the room during lunch and dinner.”

IDI, Returnee-migrant

Some others shared that their company provided them an apartment with good facilities. They did not have any complaints in-relation to their accommodation and facilities. However, they often encountered unacceptable behavior of their partners that created stressful and sometimes insecure living environments.

> “We often faced problem due to unfriendly living environment there. In the shared accommodation, some used to call home at late night, while others came late at night and knocked the door disturbing to others. Some showed their aggressive behavior and they frequently fought among themselves. We often felt disturbed and insecure as a number of friends involved in fighting.”
>
> IDI, Returnee-migrant

Non-migrant workers reported that most of them own their houses with basic facilities as per local standards, however, some of them were living in company accommodation or at construction sites in temporary sheds where they did not have basic facilities.

> “The contractor provided the accommodation. Indeed, we built very simple temporary room at work site. We used the toilet of house owner if he is living nearby. Otherwise we built simple toilet for temporary use.”
>
> IDI, non-migrant

#### Individual behavior and life style

The individual’s physical condition, life style and behaviors such as substance use were reported as factors contributing to adverse health conditions and mental illness. Participants stated that their smoking habit and alcohol use had affected their health. They also reported that alcohol was banned in some Gulf countries so they used drugs. Drug use was reported higher among those working in

Malaysia. Non-migrants also reported their drinking and smoking habits but none of them described themselves addicted.

> “Sale of alcohol and its consumption was prohibited there ……. it was strict, we had to go far from the company to consume alcohol. We didn’t have sufficient time to go there frequently for it. However, I have seen a number of friends who regularly drank. I was also addicted for a certain time.”
>
> FGD, Returnee-migrant

One of the returnee migrants stated that the individual himself is responsible for his health. He further stated that maintaining a healthy life style with regular physical exercise, good sleep and healthy diet are necessary.

> “I never experienced any health problems because I did regular physical exercises. I was previously in Nepal army and used to do exercises. ……well, people do negligence in eating, sleeping with no exercises and get sick. In fact, our health depends on ourselves. In my opinion, those who follow healthy lifestyle, they become healthy wherever they go.”
>
> IDI, Returnee-migrant

Study participants also stated that free time to go out for entertainment, relaxing and spending time with friends and families was important to remain free from mental illness and disease. However, most returnee migrants stated that they had limited free time to go out for entertainment. Long working hours with no time to relax made their livings monotonous and stressful all the time while abroad.

> “We used to work for 12 hours including over time every day. We were engaged from early morning to late evening at work. Then, we spent some times on the phone talking to the friends and families to Nepal. We sometimes spent time watching video, news, film song using mobile.”
>
> IDI, Returnee-migrant

Migrant workers reported that they often went on dating activities and engaged in sexual activities abroad. Many of them enjoyed such activities as they were away from their family and did not have any other form of entertainment. Also, they took it as one of the means to get rid of tension that they often faced due to their family problems and work stress.

> “Well, many friends spent money for girls and sex worker, playing games and finished their money and couldn’t meet their family expectation. I found this in Qatar but not possible in Saudi. Saudi is too strict. I was new for Qatar as I worked for 20 months. My friends often asked me to visit girlfriends there. “
>
> IDI, Returnee-migrant

A few non-migrant participants also reported similar stories that they worked long hours and did not have time to go out for entertainment. However, most of them stated that they did not have a very heavy work load and they spent their leisure time with family at home and also worked in their own farm together with family members.

#### Job Nature and Income Status

Returnee migrants reported that large numbers of Nepalese migrants were unskilled or semi-skilled. The majority of them were outdoor laborers and some of them had indoor cleaning jobs and also trained and skilled technical roles. The commonly reported jobs were scaffolders in construction companies, machine operators, truck drivers, cleaners, store-keepers and office jobs. Machine operators and scaffolders were reported as having the riskiest job roles where the workers often got accident and injuries. However, all returnee migrants reported monotonous job roles having long work hours without breaks and limited holidays; they often felt exploited.

> “Nepalese migrant workers usually are in labor related jobs mainly in outdoor works. Manpower Agency commits to provide company related works but some people don’t get the job as agreed. They are mainly involved in supply area. In supply sector, workers should do any works what the employers want. They don’t have fixed work and working place. They should do hard work there.”
>
> FGD, Returnee-migrant

The job nature most reported by the non-migrant participants included agriculture and construction laborers e.g. brick layer. Others reported their jobs as office assistants, security guards, IT and electrical technicians. Few of them said they worked as tailors and school teachers. However, most participants reported their work was not very hard and stressful. Both migrant and non-migrant workers stated that they had low pay jobs and their income was not enough to meet their family expenses. One returnee migrant stated that:

> “With the low level salary, it is not easy to earn as expected abroad. Family, parents and children should not assume that migrant can earn more money abroad. We always expect love and respect from the family while remaining abroad which we do not get rather family always talk about money.”
>
> IDI, Migrant

Similarly, non-migrant participants expressed that their income in Nepal was too low compared with the migrant’s income abroad and was not enough to maintain their family expenses. They showed dissatisfaction about their work conditions, low pay rate, basic working facilities, and a few of them reported that they felt stretched with multiple job roles.

> “People are working hard but paid less. My job is not so hard as compared to other [labor] jobs. It is not physically hard. It is technical skill job though I am paid less. Some people do more than one job because of low wages. They work in two to three different places and some even work for long hours. So, Nepali workers are stressful in-county as well.”
>
> IDI, Non-migrant

#### Work Environment and Employment Condition

The culture and communication in the destination country, and the work environment and working conditions were reported as major factors creating mental distress abroad. Health and safety measures applied by the company or employer, provision of insurance, health service availability when in need and also the quality of care were important factors affecting their mental health and wellbeing. Moreover, participants reported that abuse and misbehavior by supervisors and managers often created them job insecurity and immense mental distress.

> “Well, the main problem of Nepali workers is communication difficulties. Nepalese do not speak Arabic and Chinese languages. Even not confident in English as most of Nepali workers have low level of education and less educated people experience more problem. It makes the working environment stressful.”
>
> IDI, Returnee-migrant
>
> “We had to work without understanding. We could notice his aggressive face if we are making mistake. Then, we sought help from our friends to sort out. It was common for new arrival to work in that environment. It took time to be familiar with the language and people. I noticed people crying there and angriness of foreman.”
>
> IDI, Returnee-migrant

Abuse and misbehavior at work created mental distress among migrant workers. Some participants reported that their supervisors created pressure. In contrast, others said that they never faced pressure from a supervisor rather they facilitated them to sort-out problems if there were any. One study participant stated that,

> “There were verbal misbehaviors to employees. Sometimes we had the threat of job termination in the case of some mistakes. We supported 2-3 Nepali migrant workers when they had measurable situation who were expelled from the job.”
>
> FGD, Returnee-migrant
>
> “Normally the supervisors or managers didn’t give more pressure at work and they treated us politely. However, all of them were not nice all the time. Some of the supervisors gave pressure and showed their unusual behaviors such as misbehave at work.
>
> FGD, Returnee-migrant

Migrant workers were often concerned about occupational health and safety. They felt insecure with the fear of accident, injury and sickness in an adverse climate abroad. Many of them reported that their company did not provide safety training and also, no provision of basic safety measures at work often made them feel insecure at work. However, others said that their company had good provision of safety at work including healthcare facilities.

> “There was the provision of safety and security department. We didn’t have permission to do any work without getting orientation on its operating procedure. Before starting the work, orientation was given. Then, we had to begin the assigned work.”
>
> FGD, Returnee-migrant
>
> “Usually, we don’t care for personal safety related things. It is said that we should use safety related materials while doing any works. Foreign employees use such things properly. Nepali workers don’t use safety related materials.
>
> FGD, Non-migrant

Insurance, healthcare service availability and quality of care were other factors reported affecting the mental health and wellbeing of the study participants, particularly for those working abroad. A majority of returnee migrants stated that they had health insurance coverage, however some of them said they did not have insurance. Non-migrant participants reported that insurance was given only for those working in an office or for reputable companies.

> “In my case, there was no provision of insurance. It was said in the agreement that there is insurance policy but in reality there was none. If we encountered any accidents, minor medical services were provided.”
>
> FGD, Returnee-migrant
>
> “If employer is license holder, we get insurance services in accident related issues. If not, we don’t get insurance facility. It is not necessary for us having license. In the case of accidents, employer is responsible for providing insurance related things”
>
> FGD non-migrant

In-relation to the health service availability, almost all participants reported that they had easy access to good quality healthcare services abroad. They received basic services or first aid within the company if not the company took care of them and sent them to hospital. Their company paid all the treatment expenses. However, a few stated that medical facilities were not available for them and they were responsible for their own treatment. Some participants said that they faced difficulties in receiving healthcare services at night, e.g. Transport issues.

> “There was a medical clinic in every camp. Company provided paper if we needed to go for treatment in a bigger hospital if it was not possible there. For major illness, they often referred to bigger hospital or we went directly. Ambulance services were easily available.”
>
> IDI, Returnee-migrant

Despite the easy availability and accessibility of healthcare services abroad, it was reported that in the case of serious illness and major surgery companies usually did not consider sent workers back to Nepal. However, the emergency surgeries and occupational injuries were supported.

> “Hospital provides good services abroad. Workers are informed by the company about their health insurance and eligible hospital for treatment. Company does not approve for major surgery abroad. However, it is compulsory to go for accidents related treatment, for instance in fracture case.Accident related cases are reported to police and it is as like mandatory to go under treatment. In fact, health services are good abroad.”
>
> IDI, Returnee-migrant

Doctors and nurses abroad reportedly behaved kindly. With a few exceptions, healthcare services were reported as very good and of a high standard. Health providers were mostly Indians and Nepalese migrants’ workers had no problems to communicate in Hindi.

> “The quality of health services abroad is much better than Nepal. Nepal’s health service is also not bad if we only concern to health services, if not compared to abroad. Hospitals and health facilities abroad allow to serve qualified professionals only after meeting their criteria. So, high quality health services are available abroad compared to Nepal.
>
> IDI, Returnee-migrant

Non-migrant study participants reported their mixed experiences on healthcare service availability and quality of care in Nepal. A significant number of non-migrant participants commonly reported that healthcare was not easily available and the cost was not affordable for them. Some of them also reported that they faced discrimination in bigger hospitals and poor quality of care based on caste, and the economic condition of the individual. However, for those that could afford it, healthcare services in terms of availability and quality were consulted to have improved over recent years.

> “Many Nepalese people have limited access to government health services because they are poor and lack of education. They don’t have knowledge about the mental health related things. In rural areas, people cannot support those people who have mental health problems.”
>
> FGD, Non-migrant

#### Unmet expectation and economic burden

Unmet expectations, economic burdens and family related problems were other important factors that often created mental distress for both migrant and non-migrant workers. Interview participants often reported that they had high expectations for their employment abroad that not met. Moreover, any financial loan that they took to pay the agent, friends and relatives always distressed them while abroad.

> “Most migrant workers do not earn as expected by their family. Generally, they earn little with their hard work. There would not be much tension if the family back home understands husband’s hard work, income level and maintain livelihoods accordingly.”
>
> IDI, Returnee-migrant “One of my friends went to depression while I used to work in Malaysia. The salary in Malaysia was not as expected. He struggled to manage his economic balance back home as he had borrowed money from friends and relatives that he couldn’t. Finally, he went to depression. He struggled to sleep at night and we took him to hospital and was taking regular medicine.”
>
> IDI Returnee-migrant

Work stress along with family problems were reported as the leading cause of mental distress. Moreover, homesickness, family illness and unhelpful supervisors not approving leave to visit family were also important stressors reported by the returnee migrants. Such problems were not faced by non-migrant workers since they remained with their family.

> “The another reason of tension abroad is due to the illness of family at home. We do not get leave to visit and support family back home during their illness. There are a number of cases of suicidal attempt, even committed suicide cases because of not getting leave from employer abroad.”
>
> IDI, Returnee-migrant
>
> “There would be stressful long duty hours and coming back to apartment in the evening, the family argument starts. I faced problem with my wife frequently, however, I tolerated and managed. Many can’t meet the family expectation. One of my roommates had very hard time with his family since his wife eloped with someone. He was always in stress and could not concentrate at work.”
>
> IDI, Returnee-migrant

Non-migrant participants expressed their views as the reasons of mental distress were due to economic reason, unemployment and poverty. Also, they explained that the reason why migrants experienced mental distress was because they were away from their family and friends.

> “People experience more stress or tension while they work away from home. I did not experience any stress or tension as I am living with my family. I usually spend day off and also the leisure time together with my wife for agricultural activities and farming. People working away from home miss such opportunity, they miss their children and family that increases stress and tension.”
>
> IDI, Non-migrants

## Discussion

This study explored mental health and wellbeing challenges experienced by Nepali male migrant and non-migrant workers. Further, the study has attempted to understand other health-related problems they encountered and the factors enabling them to remain mentally and physically healthy. Study findings suggest that both migrant and non-migrant workers experienced mental health problems and vulnerabilities that included stress, anxiety, depressive symptoms, and exposure to potential injury and traumatic life events. However, migrant workers experienced a greater level of adverse mental and physical health problems and vulnerabilities than their non-migrant counterparts. The study also noted that those problems could potentially escalate into more serious complications leading to suicide [4]. Study findings further show that the both migrant and non-migrant workers having little knowledge about mental illness, and its long-term impact. Previous studies conducted among Indonesian [20], Myanmar [21] and cross-border Nepalese [7] migrant workers revealed the consistent findings that migrant workers are more at risk of developing adverse mental health conditions than non-migrants.

Limited knowledge about mental health can be explained as most of Nepalese labor migrant and non-migrant workers are from low educational background and they may not be fully aware about mental health problems and associated risks.

In addition to mental health conditions, study participants reported other Illness and health problems they experienced. Common illnesses encountered by both migrants and non-migrant workers were fever, seasonal flu, coughs and colds, sore-throats, and occupational injuries. Occupational health risks were more prevalent among migrants compared to non-migrant workers. Migrant workers reported more accidents and injuries due to their engagement in risky works with inadequate safety training, poor application of safety measures at work, and work pressure by the supervisors [5]. Furthermore, typical health problems faced by migrant workers included ulcers, heat rashes, urinary or kidney related problems, sexually transmitted diseases (STDs), malaria, dengue and snakebites, while non-migrants reported hypertension, diabetes and heart diseases. The findings show that migrant workers encounter acute diseases that may last for short periods, whereas the non-migrants suffer from more long term disease burden. This finding is in line with the increasing trend of non-communicable diseases among the Nepalese population [22] and also, high susceptibility to diabetes, chronic kidney and cardiovascular diseases among South Asian populations [23] and may indicate serious public health risk among the population at large.

Consistent with the findings from previous studies [24,25], this study also found that mental health problems among migrant workers persist as a result of poor living and work environments. On many occasions, migrants’ experienced their psychosocial health threatened due to communication barriers, abuse and exploitation by supervisors. In addition, long work hours without breaks and holidays, and separation from the family contributed to adverse mental health status among migrant workers. It was obvious that non-compliance of labor agreements, and violation of labor rights by the employers [26], and unaware migrant workers about their rights and entitlements such as rights of healthcare facilities, insurance and minimum wage rates were also the root causes of health vulnerability particularly among migrant workers. Amendment of inconsistent labor laws with ILO Conventions (27) by destination countries that ensure international minimum rights including ILO fundamental labor rights (28) may help to limit exploitation and abuse. It may ultimately mitigate mental distresses experienced by migrant workers.

Another important finding of this study was that the health service availability and burden of healthcare expenses while getting sick found associated with the mental well-being of both migrant and non-migrant workers. There was easy access to quality health services in destination countries for the migrant workers who were covered by insurance policies. However, consistent with the findings of other studies [7,21], this study also found that illegal migrant workers, who worked in small or disreputable companies with no insurance policy couldn’t afford the health services abroad. Despite the availability of quality health services, the employer often denied providing treatment for mental health and serious long-term illness except the case of occupational accident and injuries. Nepalese migrants are often covered with insurance only for the first year of work abroad [28].

Returnee migrant and non-migrant workers both reported that quality health services are available in Nepal as well, however, it is often not affordable for them in the case of serious illnesses. Moreover, study participants reported that they faced discrimination in bigger hospitals based on their caste, and economic status while seeking services. Past studies revealed healthcare service disparities faced by Dalits, a poor and vulnerable population in Nepal [29]. Provision of easy available health services without discrimination and financial support from employers for their medical expenses could relieve their stress related to healthcare.

This study found migrants’ adverse mental health conditions are largely rooted in their income, unmet expectations, and family problems. Usually migrant workers sought their jobs abroad with the dream of changing their life status and improving the economic condition of their family. Almost all migrants went to abroad for work through agents and in many cases, the agent/brokers gave them false information of work condition and earnings, was far below reality and their expectation so much, so that they are unable to repay loans and meet family expectations. Furthermore, sexual and emotional needs of their spouse would be unmet due the separation between husband and wife for the long duration. This often lead to relational conflict, divorce and family fragmentation creating enormous psychological distress, particularly to migrant workers. Similar findings are documented from the previous studies among other Asian migrant workers, including Nepalese migrants. [20,24,30].

### Study limitation

We acknowledge a number of study limitations. The findings are best considered as a preliminary insight into the experiences of migrant and non-migrant workers’ mental health and wellbeing as yet insufficient to support the generation of theory. The views expressed by participants in this study may not fully represent the experience of all workers as participants were male and recruited from limited countries (GCC and Malaysia) and one local unit in Nepal. This could have biased our sample towards those with more adverse experiences such as women [8] and those worked in other contexts and countries. Reliance on participants’ retrospective accounts of their experience of past illness could have led to underreporting, and also the possibility of recall bias as they may have recovered fully and forgotten the symptoms and fear related to adverse mental health and vulnerabilities. Furthermore, the current study also did not include employers’ perspectives. Another limitation is that due to the qualitative nature of this study, generalizability of the findings and establishing linkages between causal factors and health outcomes is not possible. Further research is needed to understand the shared and unique experiences of those groups.

## Conclusion

There are common mental health problems and other illnesses for both migrant and non-migrant workers, however, many differences in nature and intensity between their experiences are apparent. Migrants encountered stress largely rooted in financial, familial, living and working conditions. Study findings indicate an emerging picture of long-term health burdens with occurrences of chronic diseases such as hypertension, diabetes, and heart disease among non-migrants, while acute but serious illness among migrant workers. Further studies demonstrating an enhanced understanding on the impact of those issues may be helpful to policy planners for the development of public health interventions. It is also recommended that labor laws inconsistent with ILO conventions in the destination countries particularly in GCC countries and also in Nepal should be amended to ensure international minimum rights, including ILO fundamental labor rights that may help mitigating any adverse mental health and wellbeing for migrant and non-migrant workers.

## Data Availability

The datasets used for this study are available from the corresponding author on reasonable request.

## Declarations

### Consent for publication

Not Applicable.

### Availability of data and materials

The datasets used for this study are available from the corresponding author on reasonable request.

### Authors’ contributions

PA conceived, designed the study and obtained the funding. PA implemented the study in the field, undertook data collection mobilizing enumerators. PA and BB transcribed and translated data from Nepali to English. HRD analyzed the data, interpreted the findings and wrote the manuscript draft. PA provided his inputs for finalization of manuscript. All authors read and approved the final manuscript.

## Acknowledgements

The authors acknowledge the support and contribution of Institute of Social and Environmental Research, Nepal (ISER-N) and the colleagues who offered organizational facilities and staff time for this research. Appreciation should go to all the dedicated field workers and those who helped facilitate the field workers in the study. The authors are grateful to those who participated in the study and shared their views and personal experiences.

